# Machine Learning identification in patients with aortic valve stenosis of a myocardial miRNA signature predictive of reverse cardiac remodelling after valve replacement

**DOI:** 10.64898/2026.09.22.26363736

**Authors:** Carlos Juárez, J Francisco Nistal, Ignacio de la Torre Cubillo, Maria Calvo, J Miguel Redondo, Joaquín Bedia, Raquel García

## Abstract

**Background:** Aortic stenosis (AS) is a valvular heart disease which imposes chronic pressure overload on the left ventricle (LV). Although LV remodelling may regress following aortic valve replacement, the molecular determinants of reverse remodelling remain poorly understood. This study develops a machine learning-based model using myocardial microRNA (miRNA) expression profiles to predict LV mass normalization one year after valve replacement.

**Methods:** Consensus selection combining Random Forest and Bayesian Network analyses was applied to miRNA sequencing data from LV biopsies of 12 matched patient pairs with AS. Selected miRNAs were qPCR quantified in an extended cohort of 89 AS patients and incorporated into predictive models. Sex-stratified analyses were performed. Model robustness was assessed using bootstrap resampling and cross-validation. Candidate miRNAs and target genes were evaluated in LV from a murine model of pressure overload induced by transverse aortic constriction (TAC).

**Results:** The combined Random Forest–Bayesian Network approach identified a three-miRNA signature (miR-517a-3p, miR-122b-5p, and miR-4683) associated with LV mass normalization after valve replacement. Validation in an extensive cohort demonstrated good overall discrimination and achieved its highest performance in women (AUC >0.80). Functional analyses identified drebrin (DBN1) as a putative target of miR-517a-3p and supported its potential involvement in pressure overload-induced cardiac remodelling.

**Conclusion:** Random Forest and Bayesian Network integrated analyses enabled the identification of a myocardial miRNA signature with sex-dependent predictive value for reverse cardiac remodelling after aortic valve replacement. Functional validation supports the relevance of selected miRNAs and their downstream targets, highlighting their potential as biomarkers and candidate regulators of myocardial recovery.

**What Is New?:**

- In surgical LV myocardial biopsies from AS patients we detected, using machine learning strategies, miRnoma changes prognostic of the LV postoperative reverse remodeling completion.
- Sex stratified predictive models reveal biologically meaningful differences that could refine prognostic modelling and future therapeutic targeting in reverse cardiac remodelling.
- Low LV miR 517a 3p and high DBN1 expressions associated persistent one-year postoperative hypertrophy in patients, a pattern consistent with post transcriptional regulation that appeared also in pressure overloaded and de-loaded mice.

**What Are the Clinical Implications?:**

- Preoperative information on the post-surgical fate of LV remodelling in patients with severe AS may be important for the therapeutic decision-making process, particularly in asymptomatic patients.
- A favourable prognostic miRNA signature might be an argument to defer valve implantation (open or TAVR) in an asymptomatic patient with with severe AS and a procedural high-risk profile.
- Conversely, a worsening miRNA signature might prompt valve replacement in a well fit asymptomatic AS patient.
- The bedside availability of this information will be key for its incorporation into decision algorithms.

## 1. INTRODUCTION

Aortic stenosis (AS) imposes chronic pressure overload on the left ventricle (LV), triggering a progressive remodelling process characterized by cardiomyocyte hypertrophy and extracellular matrix accumulation. Although these structural adaptations are initially compensatory, persistent pressure overload ultimately promotes maladaptive remodelling, ventricular dysfunction, and heart failure. Relief of the valvular obstruction through surgical or transcatheter aortic valve replacement initiates a process of reverse cardiac remodelling, during which structural and functional myocardial abnormalities progressively regress (1–3). Regression of LV hypertrophy begins soon after valve replacement; however, the process extends over several months, and a substantial proportion of patients fail to achieve complete normalization of left ventricular mass despite successful correction of the valvular lesion (2,4,5). Importantly, persistent postoperative hypertrophy is independently associated with worse long-term survival and reduced functional recovery (2,6). Identifying patients with limited reverse remodelling potential before intervention therefore remains an important unmet clinical need and may facilitate more individualized therapeutic strategies.

MicroRNAs (miRNAs) have emerged as key post-transcriptional regulators of cardiac remodelling and represent promising biomarkers for disease progression and therapeutic response in (7–9) AS. Transcriptomic profiling of myocardial RNA expression has improved our understanding of the molecular mechanisms underlying pressure overload-induced remodelling and offers new opportunities for identifying biomarkers associated with reverse remodelling after valve replacement (10–12). Moreover, integrative analyses of miRNAs and their downstream targets may uncover regulatory pathways that could eventually serve as novel therapeutic targets (13,14).

The increasing availability of high-dimensional transcriptomic datasets has stimulated the application of artificial intelligence (AI) and machine learning (ML) methods in cardiovascular research. These approaches are particularly well suited to identifying complex molecular signatures, selecting informative biomarkers, and developing predictive models from datasets with a large number of variables. Random Forest, Bayesian networks, and other machine learning algorithms have shown considerable promise for patient stratification and outcome prediction across multiple cardiovascular conditions (15–19). Consequently, integrating transcriptomic profiling with machine learning represents a promising strategy for identifying molecular predictors of reverse cardiac remodelling in patients with AS.

In the present study, we combined myocardial miRNA sequencing with ensemble machine learning approaches to identify preoperative molecular signatures associated with reverse remodelling after aortic valve replacement. Specifically, we aimed to (i) identify myocardial miRNAs differentially associated with favourable and unfavourable reverse remodelling one year after valve replacement, (ii) develop and validate a predictive model for left ventricular mass normalization based on myocardial miRNA expression, and (iii) investigate candidate target genes and molecular pathways potentially involved in the regulation of reverse cardiac remodelling.

## 2. METHODS

### 2.1. Study population

The study included patients with severe aortic stenosis (AS) undergoing surgical aortic valve replacement at the Department of Cardiovascular Surgery of Hospital Universitario Marqués de Valdecilla, Santander, Spain. Patients were excluded if they had more than mild aortic or mitral regurgitation, significant coronary artery disease (>50% stenosis), previous cardiac surgery, active malignancy, or severe renal or hepatic dysfunction. Additional exclusion criteria included extreme frailty, contraindications to cardiac magnetic resonance imaging, and advanced conduction abnormalities.

During surgery, subepicardial myocardial biopsies were obtained from the lateral wall of the left ventricle (LV) using a Tru-Cut needle. The study was conducted in accordance with the Declaration of Helsinki and was approved by the Clinical Research Ethics Committee of Cantabria. All participants received written and verbal information about the study and provided written informed consent. Standardized echocardiographic examinations were performed before surgery and one year after valve replacement to assess changes in cardiac structure and function. LV mass was calculated according to the Devereux formula and indexed to height raised to the power of 2.7. LV mass normalization at one year was defined as a left ventricular mass index (LVMI) <51 g/m^2.7^.

#### 2.1.1. Discovery cohort

The discovery cohort comprised 24 patients: 12 who failed to achieve LV mass normalization one year after valve replacement (non-normalizers) and 12 who achieved LV mass normalization (normalizers). Each group included 12 men and 12 women. Normalizers and non-normalizers were matched for age, sex and preoperative degree of LV hypertrophy. Demographic and clinical features of patients are described in table S1.

#### 2.1.2. Validation cohort

The extended validation cohort comprised 89 patients. These patients fulfilled the same diagnostic and eligibility criteria, underwent the same surgical procedure, and had demographic and clinical characteristics comparable to those of the discovery cohort (table S2).

### 2.2. Sample processing and RNA isolation

Total RNA was extracted using the miRNeasy Kit (Qiagen, Germany) according to the manufacturer’s instructions. RNA quantity and integrity were assessed using an Agilent 2100 Bioanalyzer, and only samples with an RNA integrity number (RIN) >8.0 were included in the sequencing analysis.

### 2.3. miRNAs sequencing

Small RNA libraries were prepared using the TruSeq Small RNA Library Preparation Kit (Illumina). miRNA sequencing was performed by the Bioinformatics Unit of the Centro Nacional de Investigaciones Cardiovasculares (CNIC; Madrid, Spain) using an Illumina NextSeq platform.

Raw 61-nt sequencing reads were subjected to quality control using FastQC (20). Adapter sequences were removed using Cutadapt (21), and reads shorter than 15 nt or longer than 35 nt after trimming were excluded to enrich for miRNA-derived sequences. The remaining reads were aligned to human mature miRNA sequences obtained from miRBase release 22, and expression estimates were generated using RSEM (22).

Expected counts were processed in R using the Bioconductor package limma (23). Library-size normalization was performed using the trimmed mean of M-values (TMM) method. Only miRNAs reaching an expression threshold of at least 10 counts per million (CPM) in the minimum number of samples predefined for each comparison were retained for downstream analyses.

### 2.4. Quantification of myocardial miRNA and mRNA expression

Expression of miR-122b-5p, miR-517a-3p, miR-4683, and drebrin 1 (DBN1) was quantified by real-time quantitative PCR using TaqMan assays (Thermo Fisher Scientific). RNU6b and 18S were used as endogenous reference controls for miRNA and mRNA expression, respectively.

### 2.5. Identification of target genes of the miRNAs of interest

*In silico* analysis of potential target genes was performed for the miRNAs identified as miRNAs of interest due to their predictive ability for left ventricular mass normalization one year after valve replacement. The following bioinformatic databases designed to identify miRNA–mRNA interactions were used: miRWalk, miRDB and miRTarBase (24,25).

### 2.6. Machine learning analysis

Given the limited sample size (24 patients), overfitting and lack of robustness posed a significant methodological concern. To mitigate this, we adopted two complementary strategies: first, the use of Bayesian networks (BNs) allowed us to incorporate probabilistic reasoning and conditional dependencies, which are particularly advantageous in small-sample contexts where data-driven inference alone may be unreliable. BNs can incorporate accumulated expert knowledge in situations where data are limited, and still produce useful decision-support systems (16,26), and have shown particular utility for learning complex dependency structures in high-dimensional, low-sample-size biological datasets (27). Second, Random Forests were implemented using bootstrap aggregation (bagging), whereby each tree is trained on a bootstrap sample of the data. This strategy reduces variance, improves generalizability, and provides robust variable-importance measures (28). By leveraging both approaches, we aimed not only to balance interpretability and robustness, but more importantly, to identify the most relevant miRNAs associated with post-surgical LV mass normalization.

#### 2.6.1. Bayesian networks

Bayesian Networks (BNs) are probabilistic graphical models that represent conditional dependencies among variables through a Directed Acyclic Graph (DAG) to represent stochastic dependencies, which are quantified through probability distributions (29–31). Formally, Bayesian Networks consist of two main components (32):

- Network structure: A DAG *G* = (*V*, *A*), where each node *v_i_* ∈ *V*corresponds to a random variable *Xi* (e.g.: a certain biomarker expression).
- Global Probability Distribution. A distribution *P*(*X*)parametrized by *Θ*, which can be factorized into smaller local probability distributions based on the arcs *a_ij_* ∈ *A* present in the graph

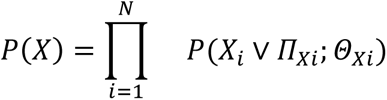

where *Π_Xi_* represents the set of parents of *X_i_*. In this context, a “parent” is a node that has a direct arrow pointing into the node of *X_i_*. Similarly, a “child” of a node is any node that has a direct arrow pointing out from that node into it. Children represent variables whose conditional distribution is directly influenced by the parent node.

In biomedical settings with limited samples and correlated biomarkers, BNs provide: (i) principled regularization via structure scores that penalize overly complex graphs, (ii) explainable inference for clinical outcomes, and (iii) a natural way to integrate data-driven evidence with domain knowledge. As a result, they have been successfully applied to biomarker discovery and to elucidate complex interactions (16,33–35).

#### 2.6.1.1. Variables and node definitions

All variables are modelled as ordinary BN nodes, with no prior hierarchy imposed; their ordering and dependency structure is determined solely by the data through structure learning.

- *miRNA nodes*. Each node represents the preoperative expression level of a candidate miRNA measured in LV myocardium. To enhance robustness and clinical interpretability, continuous expression values are discretized into binary categories (“low”/“high”), as detailed below.
- *Outcome node* (“norm”). Normalization is encoded as a binary variable (“yes”/”no”) indicating LVMI normalization at 1 year (< 51 g/m^2.7). During structure learning, the normalization node is treated as any other node in the network, with no constraints imposed on the direction of incoming or outgoing edges. Following network construction, analyses are focused on the Markov Blanket of the normalization node and on the network’s ability to predict this outcome, allowing identification of the variables most strongly associated with post-surgical normalization while retaining a fully data-driven structure learning process.
- *Optional clinical/stratification nodes*: variables such as sex, age, or baseline echocardiographic indices may be included to adjust for confounding and to enable subgroup analyses (e.g., sex-specific models). Including such nodes helps parse whether a miRNA acts directly on the outcome or through clinical intermediates.

##### 2.6.1.2. Encoding miRNA expression for BN construction

The miRNA expression dataset was pre-processed before model construction following two key steps:

1. Preprocessing and filtering: normalized count data (e.g., TMM-normalized counts) are filtered to retain miRNAs with sufficient expression and variability to avoid spurious signals.
2. Supervised discretization: continuous miRNA values are discretized into binary states using Hartemink’s iterative information-theoretic method, which optimizes cut-points jointly across all variables to improve the global Bayesian network score (36). This preserves predictive structure relevant for probabilistic modeling, reduces variance under small *n*, and yields clinically interpretable categories (“low” vs. “high”).

Following preprocessing, each discretized miRNA was incorporated into the Bayesian network as a categorical node. The learned network structure determines whether a miRNA is associated with Normalization directly or through interactions with other variables.

##### 2.6.1.3. Structural Learning and model evaluation

Bayesian network structure learning aims to identify the Directed Acyclic Graph (DAG) that best represents the dependencies among variables while balancing goodness-of-fit and model complexity. To compare alternative structure-learning algorithms, we performed 10-fold cross-validation with 10 repetitions. Two loss functions were evaluated: log-likelihood (logLik), which measures the ability of the learned network to explain unseen data, and classification error, which quantifies predictive performance for a target node based on its local distribution (i.e., its parents). The target node was norm, representing post-surgical LV mass normalization. Score-based algorithms, particularly hill-climbing and tabu search, consistently achieved the lowest classification errors (Fig. S2), indicating superior performance for modeling normalization outcomes. Based on these results, tabu search was selected for subsequent analyses. Further details are provided in Fig S2 and the corresponding description in the section *structural learning method selection experiment* in Supplementary Material. Based on this evidence, we selected the *tabu search* algorithm for final network construction (37–39). These findings are consistent with previous studies showing that score-based Bayesian network algorithms are particularly effective at recovering complex dependency structures in high-dimensional biological datasets (27,40).

##### 2.6.1.4. Markov Blanket based approach for key miRNA identification

Edges (arcs) in the DAG encode probabilistic dependencies among miRNAs and between miRNAs and norm; an incoming edge into norm indicates that the parent adds information about normalization probability conditional on other parents. The Markov blanket (MB) of a node—the union of its parents, children, and the parents of its children—is the minimal set that renders the node conditionally independent of all remaining variables (41). Thus, inference about norm can be based solely on its MB. In our context, focusing on the MB of the node-*norm* isolates a minimal, most informative subset of miRNAs with direct predictive or mechanistic relevance, making this approach especially suitable for high-dimensional, low-sample biomarker discovery (42,43).

Given the high dimensionality (239 miRNAs) relative to sample size, learning a full BN over all variables would yield unstable structures and parameters. To enhance robustness, we implemented a bootstrap-MB frequency strategy (44): we generated 100 bootstrap replicates, learned a BN with tabu search in each replicate, and extracted the MB of norm (Fig. S3). MiRNAs were ranked by their frequency of occurrence in the MB across replicates, highlighting variables with stable, meaningful associations to the outcome and prioritizing candidates for a compact, actionable biomarker panel.

#### 2.6.2. Random forests

Random Forests (RFs) are ensemble classifiers introduced by Breiman (2001) (28). They construct a collection of decision trees, each trained on a bootstrap sample of the data, and classify new observations by majority voting across trees. Given a training set with *N* samples and *M* predictors, each tree is built by drawing *N* observations with replacement (bootstrap sampling). At each internal node, a random subset of *m* predictors is selected, and the best binary split among these *m* features (typically evaluated using the Gini impurity) is applied. This process continues until a minimum terminal node size is reached, and predictions for new samples are obtained by aggregating votes across all trees (28,45).

To ensure methodological consistency and enable direct comparison with the Bayesian network analysis, we used the same variables and the same discretized miRNA expression dataset described in the previous section. Although Random Forests can naturally accommodate continuous predictors, we intentionally maintained the discretized representation to ensure that both modelling frameworks operated on identical data transformations. This facilitates a direct cross-method validation of candidate miRNAs and enhances the comparability of variable-importance metrics between BN and RF analyses.

Hyperparameters known to influence RF performance were optimized empirically. The final model used 1,000 trees, 15 randomly selected variables considered at each split (*m*=15), and a minimum terminal node size of 3.

Model discrimination was evaluated using the Area Under the Receiver Operating Characteristic Curve (AUC-ROC) as the primary optimization criterion. To assess generalizability, we performed stratified 5-fold cross-validation, preserving the 50/50 class balance within each fold. Performance metrics were averaged across folds to obtain robust estimates and corresponding confidence intervals.

#### 2.6.3. Variable importance assessment

To identify the most robust and relevant miRNAs in Bayesian Networks, we developed a Reduced Markov Blanket methodology that combines bootstrap stability assessment with co-occurrence network analysis. Following the extraction of Markov blankets for the target variable (’norm’) across the 100 bootstrap iterations, we applied a frequency-based filtering approach to retain only variables appearing in a predefined proportion of bootstrap samples, as a balance between robustness and biological relevance (Fig. S3). To further characterize their joint stability, we constructed a symmetric co-occurrence matrix quantifying how often pairs of miRNAs appeared together in the Markov Blanket across bootstrap samples. This matrix summarizes recurrent dependency patterns and supports the identification of miRNAs that consistently participate in the local dependency structure of the outcome.

Variable importance in Random Forest was assessed using two complementary RF metrics: Mean Decrease Accuracy (MDA) and Mean Decrease Gini (MDG). MDG measures the cumulative decrease in Gini impurity attributed to each feature across all trees, reflecting its ability to produce purer, more homogeneous splits (46). Higher MDG values indicate stronger individual discriminative power. MDA evaluates importance by permuting each variable and quantifying the resulting drop in predictive accuracy (28). This permutation-based metric captures variable interactions and nonlinear contributions that may not be fully reflected in split-based measures (45,47). Together, MDG characterizes individual splitting strength, whereas MDA captures overall predictive contribution within the ensemble. miRNAs ranking highly in both metrics were designated “consensus miRNAs”, representing robust candidates with stable discriminative and predictive relevance across complementary importance criteria.

##### 2.6.3.1. Consensus Score Calculation

To achieve robust miRNA identification, we integrated importance rankings from Bayesian Networks and Random Forest. To this aim, we calculated a Standardized Importance Score allowing to effectively intercompare the importance of miRNAs as measured by different methodologies. Both RF combined scores (sum of standardized Mean Decrease Accuracy and Gini) and BN scores (Markov blanket frequency percentage) were *z-score* standardized to enable direct comparison across methods, that is: *z* = (*x* − *μ*)⁄*σ* where *x* is the raw score, *μ* is the mean, and *σ* is the standard deviation. Hence, the consensus score for each variable is calculated as:

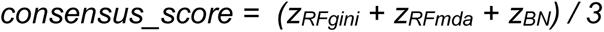

where *zRFgini*, *zRFmda and zBN* are the standardized scores of miRNAs in the Random Forest model (Gini and Mean Decrease accuracy) and the BN model (frequency of appearance in Markov Blanket). miRNAs were categorized as “Both Methods” (detected by RF and BN), “Random Forest Only”, or “Bayesian Network Only” based on their presence in each method’s variable set. However, only miRNAS appearing in all methods were subsequently considered of interest and are presented in the consensus analysis results.

#### 2.6.4. Validation and Predictive Modeling

In the second phase of the study, we aimed to independently validate the relevance of the miRNAs identified in the discovery phase using quantitative PCR (qPCR) data. The validation cohort comprised 89 LV biopsies from AS patients undergoing valve replacement. The cohort included 44 male and 45 female patients, with a balanced distribution of cardiac function normalization outcomes (47% non-normalizers, 53% normalizers). The qPCR data were then used, in combination with sex information, to develop predictive models of post-surgical recovery.

Predictive models were developed using supervised machine learning techniques, namely Random Forests (RF), Logistic Regression (Generalized linear model with binomial family for interpretable coefficient estimation, LR) and Naive Bayes (NB), a probabilistic classifier assuming feature independence, as previously checked. Naive Bayes was selected for the final validation phase as a natural extension of the Bayesian framework used in the exploratory analysis, where BNs were applied to uncover complex dependencies among a larger set of variables. This classifier is equivalent to a Bayesian network with a star-shaped structure, where the target variable is at the center and all other variables are directly connected to it (48). This simplification makes it particularly effective and computationally efficient when working with a small number of features. Since the final validation stage focused on a reduced subset of previously selected miRNAs, the use of Naive Bayes allowed us to build interpretable models with minimal risk of overfitting.

The use of cross-validation as our primary validation strategy was driven by sample size constraints in stratified analyses. While test set validation remains as a standard for large datasets, cross-validation provides more robust estimates for limited samples and reduces the risk of overfitting that can occur with small test sets. Hence, performance was evaluated using 10-fold cross-validation to ensure robust and generalizable estimates (49). Model performance was assessed using Area Under the Receiver Operating Characteristic Curve (AUC-ROC), Sensitivity and specificity from confusion matrices, with cross-validated performance estimates with 95% confidence intervals. All models used exclusively miR-122b-5p, miR-517a-3p, and miR-4683 as predictors for normalization status, with *sex* used for stratification but not as an explicit predictor variable.

Although *sex* was included as a covariate in the initial modeling phase using Bayesian networks and random forests, no significant sex-specific patterns were detected at that stage (being ‘sex’ not ranked among the most influential variables) likely due to the limited sample size. Nevertheless, given the known sex differences in cardiac remodelling in AS patients, separate models were developed for male and female subgroups using identical methodological approaches. Henece, sex stratification was performed *a priori* based on biological rationale rather than *post-hoc* data exploration.

##### 2.6.4.1. Variable importance assessment in validation models

While the consensus analysis prioritized miRNAs based on their cross-method consistency, variable importance assessment in predictive models serves a complementary purpose by quantifying each biomarker’s direct contribution to accurate prediction of normalization outcomes. For Random Forest, variable importance was quantified using the Mean Decrease in Accuracy metric derived from out-of-bag samples. Hyperparameter optimization was performed using 5-fold cross-validation by evaluating alternative m values (the number of predictors randomly considered at each split). The value maximizing the mean cross-validated classification accuracy was subsequently used to fit the final forest of 1,000 trees. Logistic Regression importance was determined by the absolute values of standardized regression coefficients, with statistical significance assessed at α = 0.05. Naive Bayes importance was calculated using permutation-based methodology, measuring the mean decrease in classification accuracy when each variable was randomly permuted across 30 iterations. All importance scores were normalized to a 0-100 scale for cross-model comparability, and visualized as a heatmap showing the complete matrix of sex-model-biomarker interactions. Sex-stratified analyses were performed separately for male (*n*=44) and female (*n*=45) patients to investigate potential sex-dependent difference patterns. Model performance was evaluated using 10-fold cross-validation with reproducible random seeds to ensure consistent results across all analyses.

All analyses were performed in R (50) using the following packages: *caret* (51) for machine learning pipelines, *randomForest* (52) for ensemble methods, *bnlearn* (41) for Bayesian Network construction and analysis, *e1071* (53). Naive Bayes implementation, *pROC* (54) for ROC analysis and *lattice* (55), *ggplot2* (*56*) and *pheatmap* (57) for visualization.

### 2.7. Transverse aortic constriction (TAC) and release (de-TAC)

All procedures were approved by the University of Cantabria institutional committee for animal experimentation (PI-03-23) and complied with Directive 2010/63/EU of the European Parliament. Experiments were performed in male and female C57BL/6 mice aged 16–20 weeks. Animals were housed under a 12-hour light/dark cycle with ad libitum access to food and water. The number of animals per group was the minimum necessary to achieve statistical significance differences (n=5-6). Groups of 4-5 mice per cage were established before surgery and individually after surgery. Animals were randomly assigned to transverse aortic constriction (TAC) or sham surgery. TAC was performed using the customized technique described by Merino et al.(58) which produces comparable degrees of pressure overload across animals of different body sizes and thereby facilitates comparisons between sexes. Animals were followed for four weeks after TAC. In a separate cohort, the constriction was subsequently released (de-TAC), and animals were followed for an additional week (58, 59).

At the experimental endpoint, mice were euthanized by exsanguination under ketamine–xylazine anaesthesia. Hearts were rapidly excised, the LV was dissected and weighed, and myocardial tissue was snap-frozen in liquid nitrogen for subsequent molecular analyses.

### 2.8. Western Blotting

Protein lysates (30 μg) were separated by 10% SDS-polyacrylamide gel electrophoresis and transferred to polyvinylidene difluoride membranes. Membranes were incubated with primary antibodies against DBN1 (Cell Signaling Technology, Danvers, MA, USA) and GAPDH (Santa Cruz Biotechnology, Dallas, TX, USA), followed by the corresponding secondary antibodies. Protein bands were visualized by infrared fluorescence using an Odyssey imaging system (LI-COR Biosciences, Lincoln, NE, USA).

DBN1 expression was quantified by densitometry and normalized to GAPDH. Samples from two mice per group were analyzed in two independent experiments.

### 2.9. Statistics

Data are expreed as means ± SEM. Differences between two independent groups were assessed with two-tailed Student’s t-test. Differences between multiple groups analysing one independent variable, were analyzed with one-way ANOVA followed by Bonferroni post hoc test. Correlations were performed using Pearson’s correlation analysis. The significance level was p < 0.05. GraphPad Prism 5 (GraphPad Inc, CA, USA) software was used for statistical analysis.

## 3. RESULTS

### 3.1. Bayesian Network analysis

The sequencing dataset was complete, with no missing values, and presented as continuous variables. Given the high number of predictors and the limited sample size, to address redundancy caused by high correlation between miRNA expression reads, we looked for pairs of variables with strong correlation (Fig. S1 and Table S3), regardless of the sign. Specifically, one miRNA from each pair with a correlation coefficient (Spearman’s rho > 0.85) was removed. Although this is a relatively high threshold, it was deliberately chosen to preserve potentially informative variables while still mitigating multicollinearity. This conservative criterion ensures that only strongly redundant signals are excluded, minimizing the risk of discarding biologically relevant information. In each correlated pair, the miRNA with lower overall relevance, based on its frequency of appearance in the Markov Blanket and its importance ranking in Random Forest models in preliminary analyses with the full set, was removed to retain the most informative candidate. These procedures reduced the initial raw set of 539 predictors to the final set of 239 miRNAs finally considered in the study, decreasing multicollinearity and improving the efficiency of subsequent models.

The frequencies of variable appearances in the resulting Markov Blankets are listed in Table 1 (See also Fig. S3 for a graphical summary). The analysis identified two core miRNAs with high stability: miR-122b-5p (39% frequency) and miR-4683 (38% freq.), along with 13 additional frequent variables (freq. ≥ 10%) including miR-4473 (19% freq.) and miR-517a-3p (17% freq.).

**Table 1.**
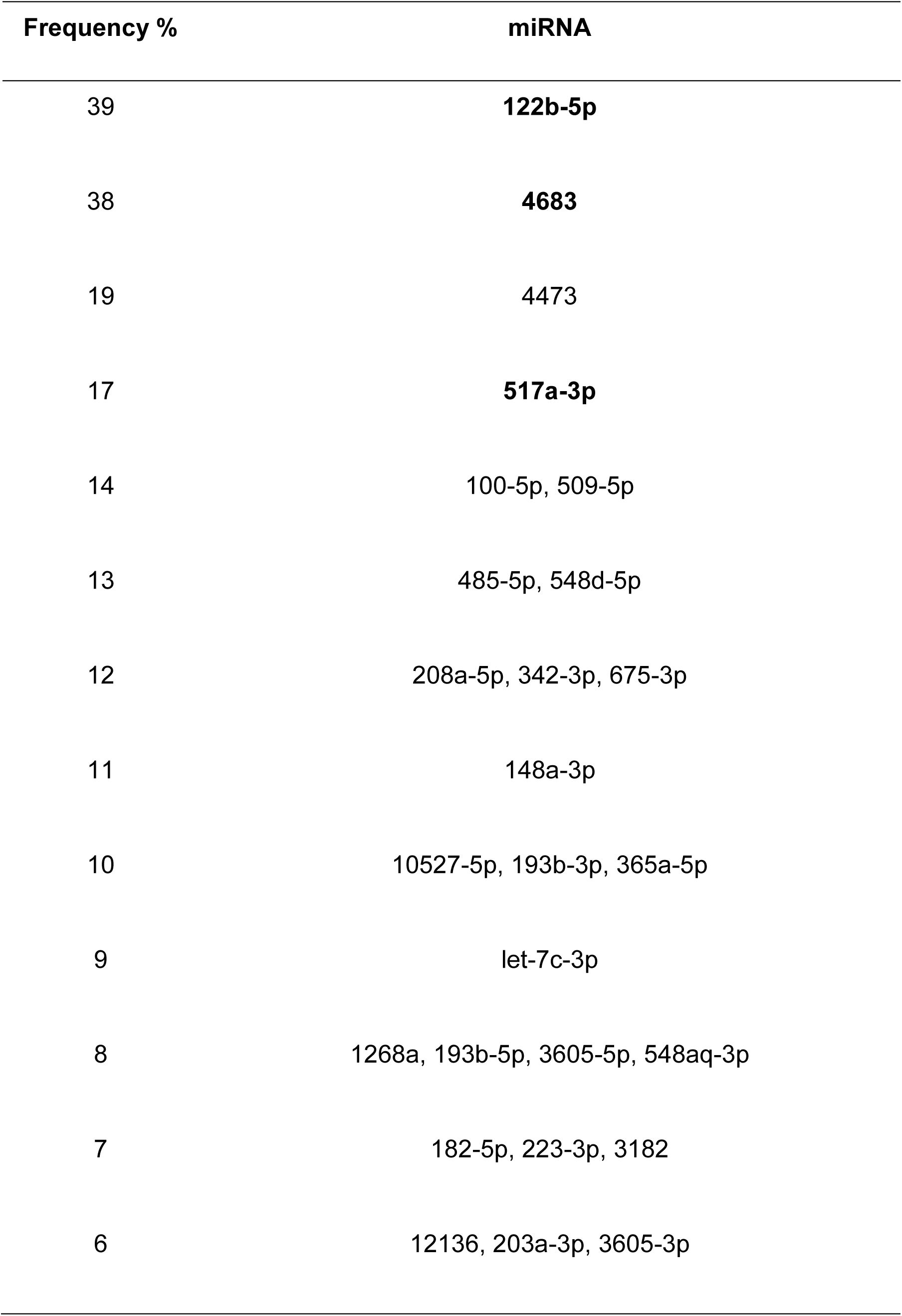
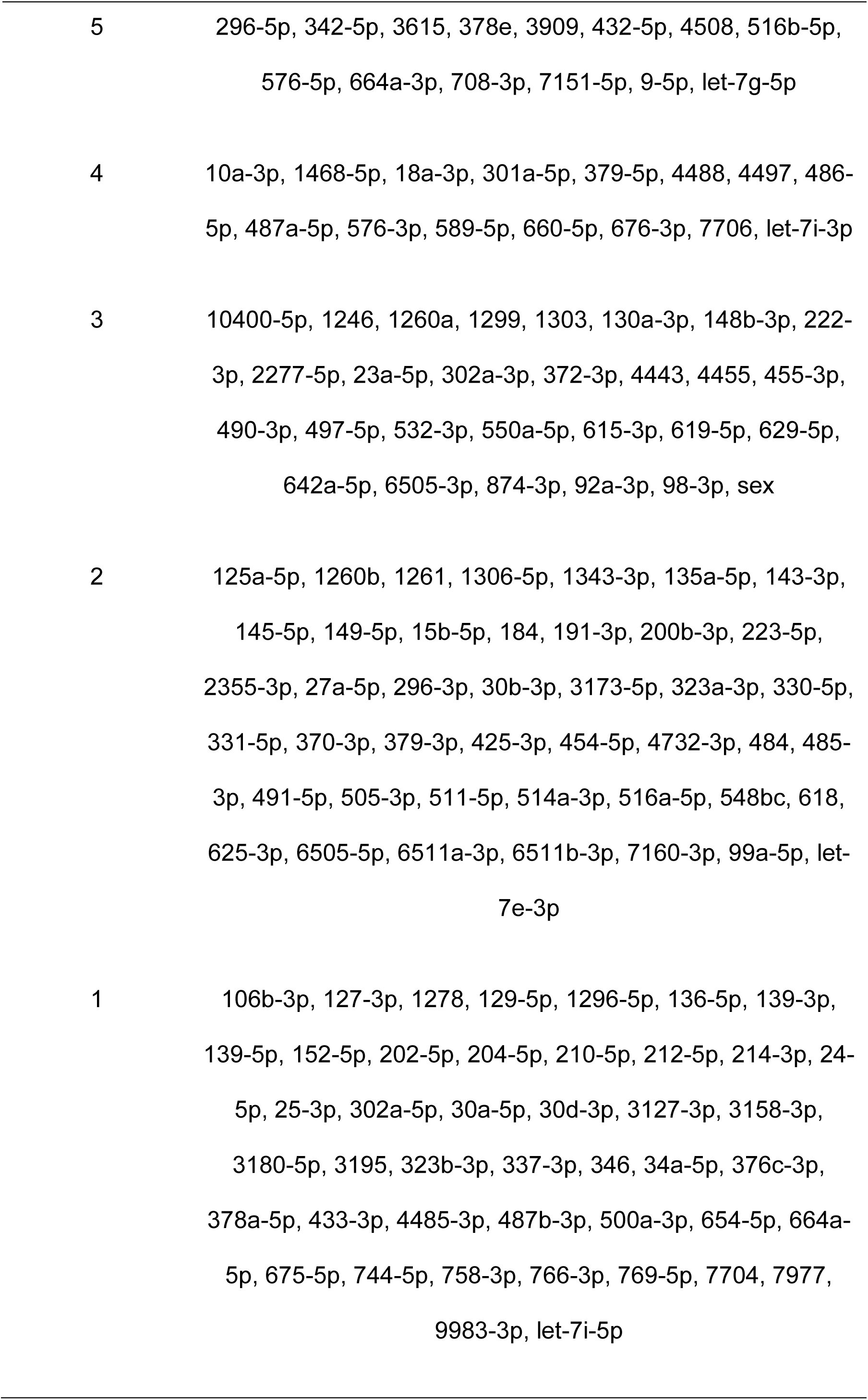
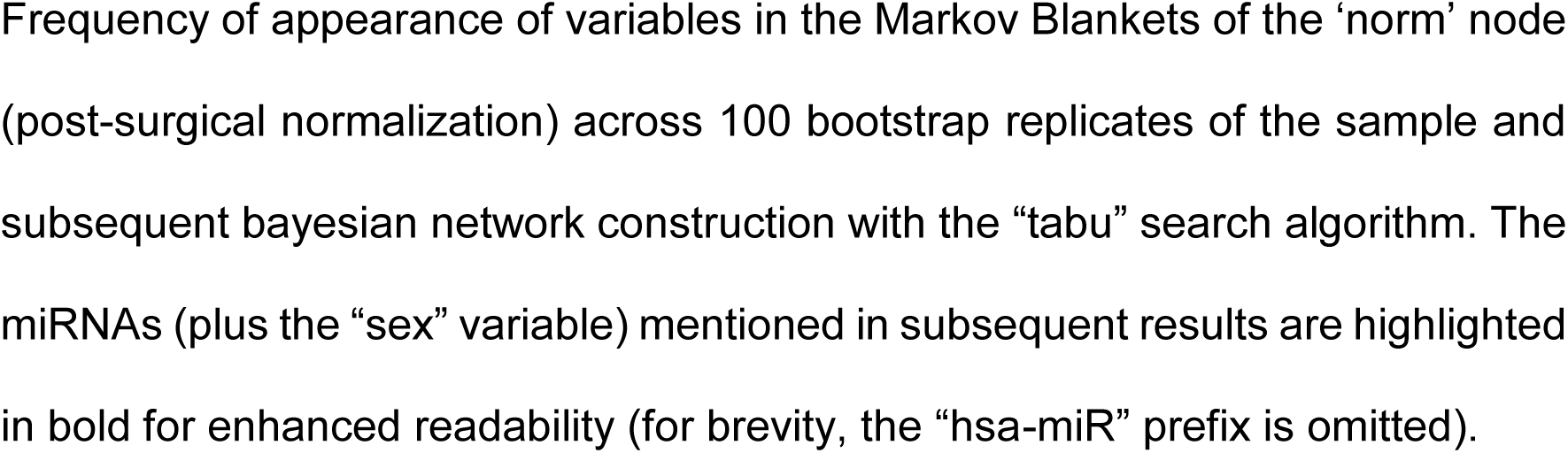
Frequency of appearance of variables in the Markov Blankets.

After calculating the reduced Markov Blanket network, here we present a visualization that allows for a quick inspection of the overall results. The network visualization employs node sizes proportional to bootstrap frequency and edge weights reflecting co-occurrence strength, providing an interpretable representation of the most stable miRNA interactions. This approach effectively reduces the dimensionality from 239 miRNAs to a focused set of ∼16 miRNAs while preserving the most consistent biological relationships, addressing the challenge of high-dimensional biomarker discovery in limited sample size in this study and allowing for visual inspection of the results. The results are depicted in Fig. 1, highlighting intricate regulatory networks involved in normalization, with most relevant biomarkers emerging as central predictive nodes.

**Figure 1.**
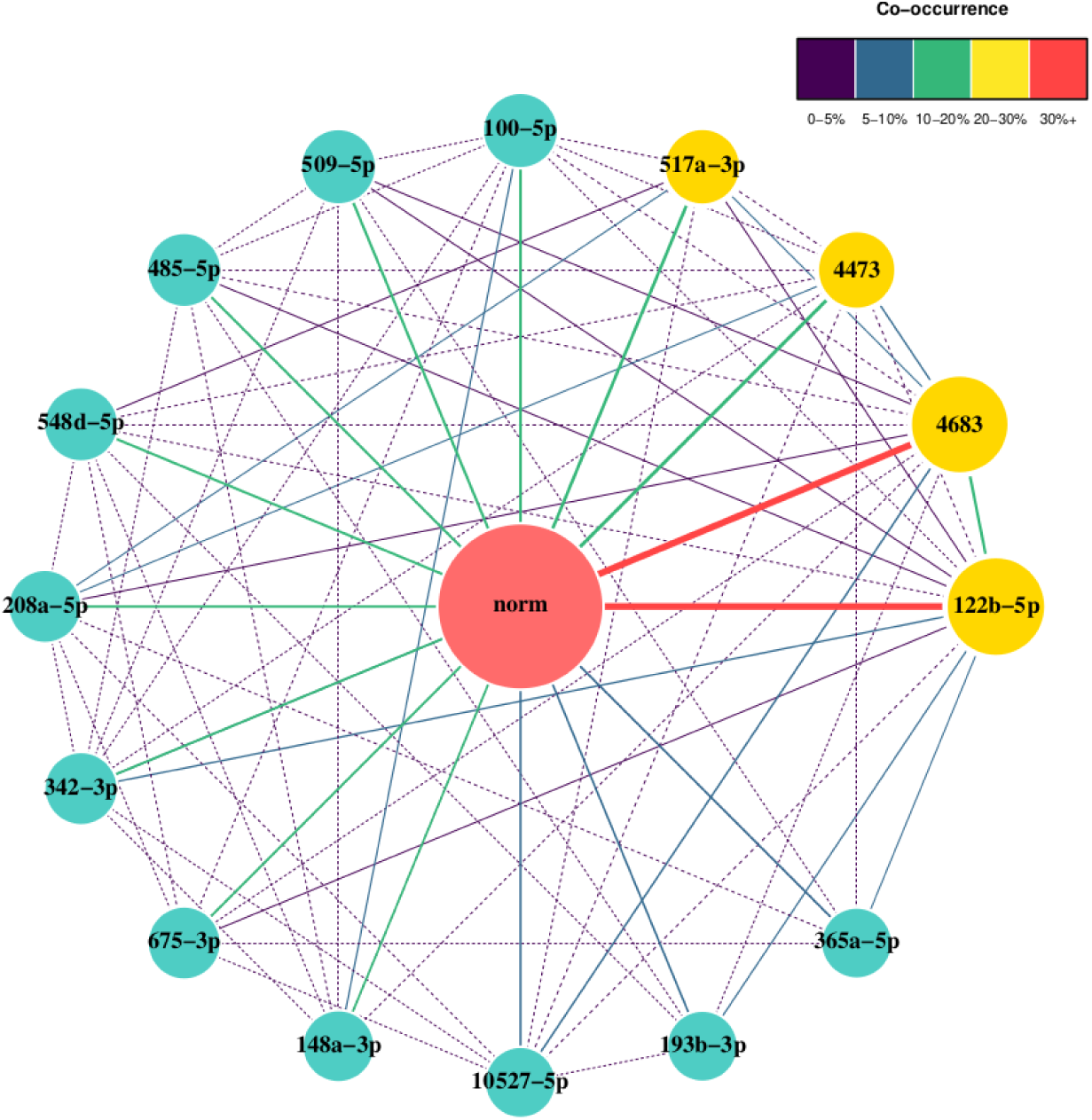
Reduced Markov Blanket Graph of miRNA-normalization associations from bootstrap Markov blanket analysis. The target outcome (“norm”) is centered, surrounded by 15 significant miRNAs (≥10% bootstrap frequency). Node size reflects appearance frequency; edge thickness indicates co-occurrence strength, also indicated by the colorbar. Solid lines represent strong associations (≥5% co-occurrence); dashed lines indicate weaker relationships (<5%). Gold nodes highlight the top 4 miRNAs most frequent in the Markov Blanket after 100 bootstrap iterations: miR-122b-5p (39%), miR-4683 (38%), miR-4473 (19%), and miR-517a-3p (17%). Light blue nodes represent additional predictive miRNAs.

### 3.2. Random Forest Analysis

Random Forest analysis provided an independent assessment of miRNA relevance. The highest-ranking variables according to both Mean Decrease Accuracy and Mean Decrease Gini were miR-4683, miR-122b-5p and miR-517a-3p, with consistent rankings across both importance metrics (Table 2; see also Fig. S4). These findings showed strong agreement with the Bayesian Network results and motivated the subsequent consensus analysis.

**Table 2:**
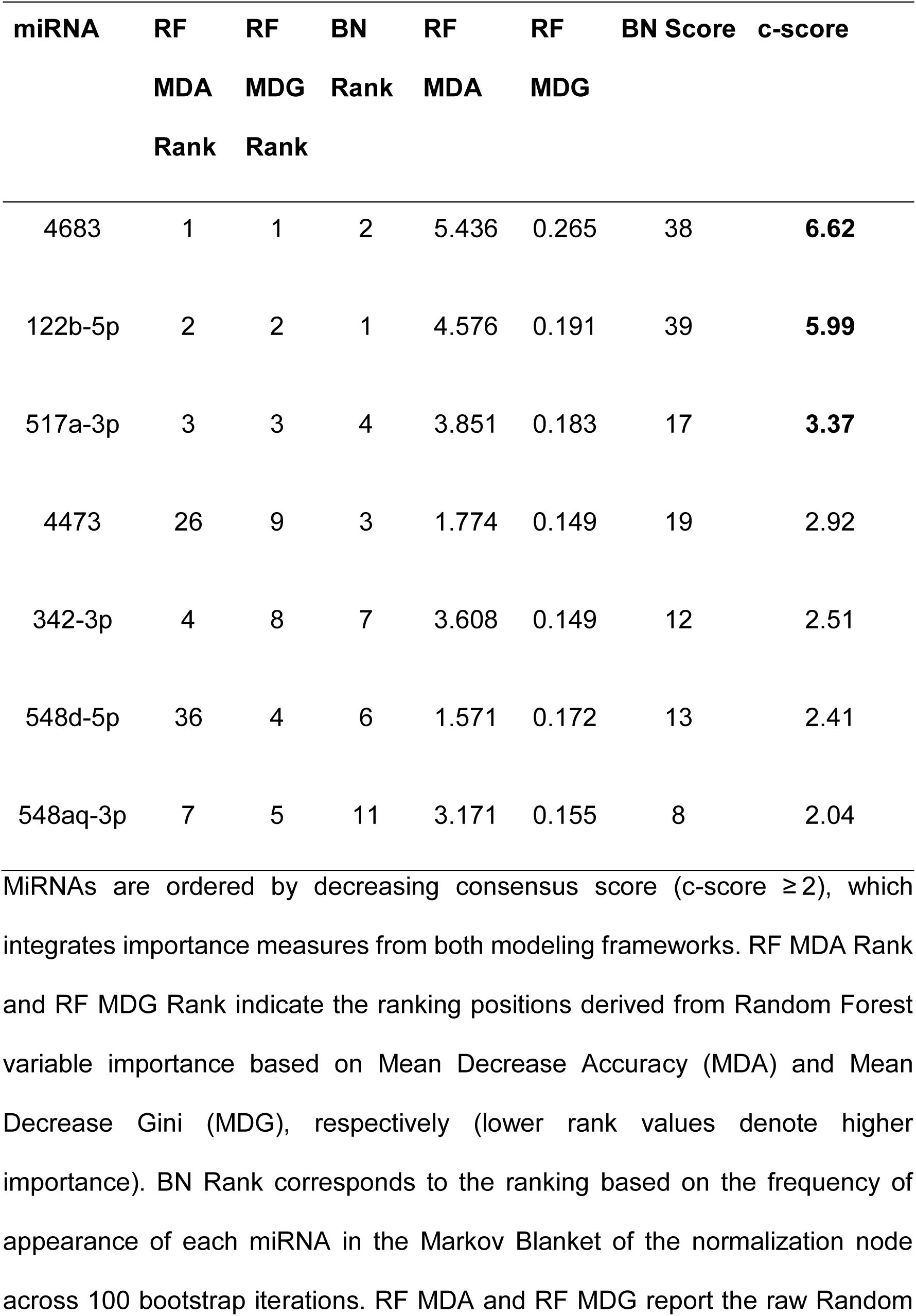
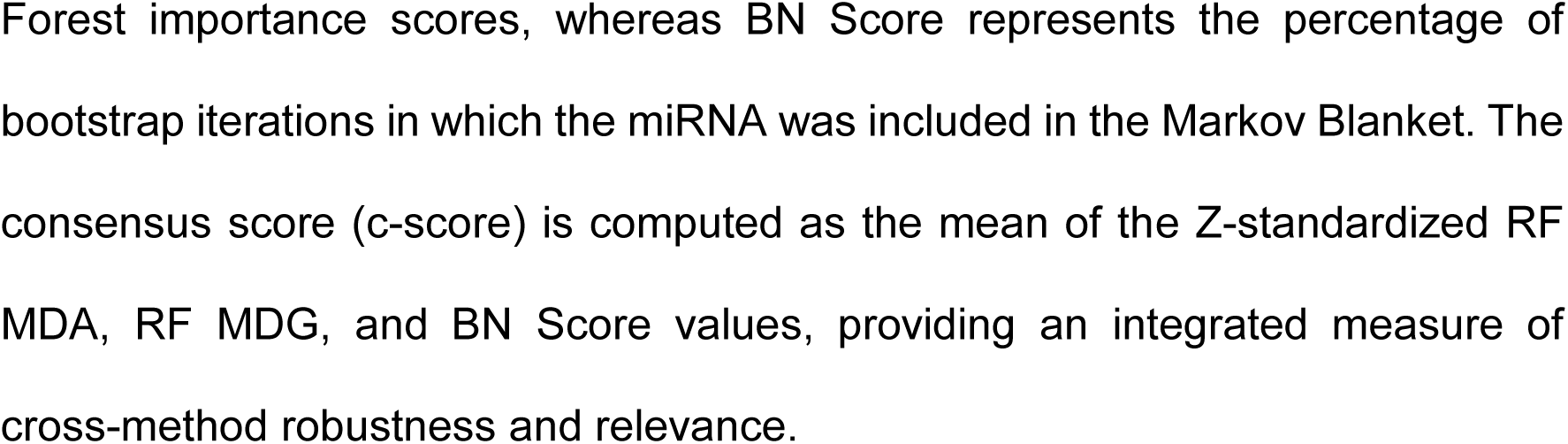
Consensus ranking of miRNA biomarkers identified by Random Forest (RF) and Bayesian Network (BN) analyses.

### 3.3. Multimodel Consensus Analysis

Key biomarkers identified by RF algorithm include miR-4683 (in first place), miR-122b-5p, and miR-517a-3p among the top-ranking variables, the three consistently following the same ranking using both Gini and Accuracy as importance measures (see also Fig. S4). Building on these results, the integrative consensus analysis combining Random Forest and Bayesian Network approaches successfully identified 80 miRNAs that were consistently detected by both methodologies, providing robust evidence for their potential regulatory roles in the normalization process (Table S4). Among these consensus candidates, miR-4683, miR-122b-5p, and miR-517a-3p emerge as the most promising targets, achieving the highest consensus scores (6.6, 6.0, and 3.4, respectively, Table 2) and demonstrating complementary ranking patterns across both machine learning frameworks (Fig. 2). Notably, miRNA-4683 ranked first in Random Forest importance and second in Bayesian Network frequency, while miR-122b-5p showed the inverse pattern (RF rank 2, BN rank 1), indicating these molecules are consistently identified as critical regulators regardless of the analytical approach. The third candidate, miR-517a-3p, maintained high rankings in both methods (RF rank 3, BN rank 4), further supporting its regulatory significance. The third-ranked miRNA identified by Bayesian Networks (BN), miRNA 4473, exhibits a substantially lower c-score (2.9), indicating a weaker association with normalization. Its importance is also markedly reduced in the Random Forest (RF) analysis, particularly in the Mean Decrease Accuracy (MDA) ranking, where it ranks 26th. This suggests that its relevance is considerably lower than that of the top two miRNAs. Therefore, the first three miRNAs represent the most statistically robust candidates for experimental validation, as they consistently demonstrate high importance across distinct algorithmic approaches and show strong associations with the normalization phenotype through both feature selection (RF) and probabilistic network analysis (BN). Accordingly, additional samples were analyzed to validate these top candidates.

**Figure 2.**
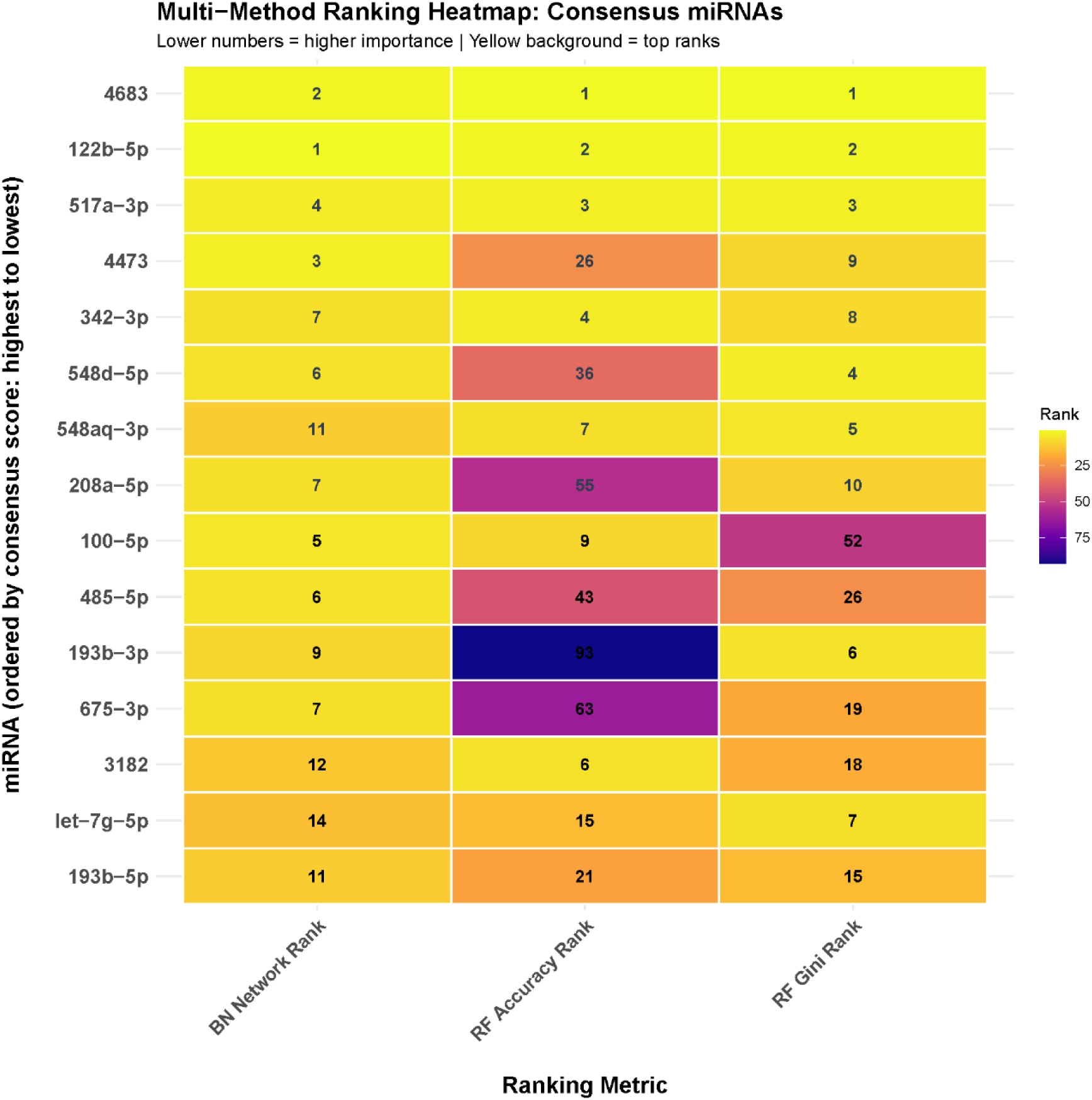
Comparative ranking visualization of the top 15 consensus miRNAs detected by both Bayesian Network and Random Forest approaches. Each row represents a miRNA ordered by consensus score (highest to lowest from top to bottom). Columns show rankings from three complementary metrics, namely BN Network Rank (Bayesian Network Markov blanket frequency), RF Accuracy Rank (Random Forest Mean Decrease Accuracy), RF Gini Rank (Random Forest Mean Decrease Gini). Color intensity represents ranking position (yellow = top ranks, blue = lower ranks). Lower numerical values indicate higher importance.

### 3.4. miRNA Validation and predictive model assessment

#### 3.4.1. Global Model Performance

The global models, trained on the complete dataset (*n*=89), demonstrated variable performance across different machine learning approaches (Table 3). Naive Bayes achieved the highest discriminatory performance with an AUC of 0.714 (95% CI: 0.575-0.853), representing good predictive capability. Random Forest showed moderate performance with an AUC of 0.686 (95% CI: 0.544-0.828), while Logistic Regression achieved an AUC of 0.627 (95% CI: 0.512-0.743). All models demonstrated fair to good discriminatory ability (AUC > 0.6), indicating that the consensus miRNA biomarkers possess meaningful predictive value for clinical outcomes.

**Table 3:** Sex-stratified 10-fold cross-validated classification performance summary of Random Forest (RF), Naive Bayes (NB) and Logistic Regression (LR) of post-surgical left ventricle mass normalization.

| Analysis Type<br>(sample size) | Model | AUC | 95% CI | Performance<br>category |
| --- | --- | --- | --- | --- |
| Global Models<br>(n=89) | RF | 0.686 | (0.544-0.828) | Fair (0.6-0.7) |
|  | LR | 0.627 | (0.512-0.743) | Fair (0.6-0.7) |
|  | NB | 0.714 | (0.575-0.853) | Good (0.7-0.8) |
| Men-Specific Models<br>(n=44) | RF | 0.517 | (0.315-0.719) | Poor (<0.6) |
|  | LR | 0.508 | (0.304-0.713) | Poor (<0.6) |
|  | NB | 0.617 | (0.446-0.787) | Fair (0.6-0.7) |
| Women-Specific<br>Models<br>(n=45) | RF | 0.775 | (0.574-0.976) | Good (0.7-0.8) |
|  | LR | 0.833 | (0.696-0.971) | Excellent (0.8-0.9) |
|  | NB | 0.842 | (0.672-1.000) | Excellent (0.8-0.9) |

hsa-miR-122b-5p, hsa-miR-517a-3p and hsa-miR-4683 miRNA expression values were used as predictors. Qualitative performance categories are according to Hosmer and Lemeshow (2000). 95% confidence intervals calculated as $mean\ AUC \pm 1.96 \times standard\ error$ , where mean AUC is the mean of the k=10 folds.

#### 3.4.2. Sex-Stratified Model Performance

A remarkable finding was the substantial improvement in predictive performance when models were stratified by sex (Fig. 3). Female-specific models (*n*=45) demonstrated excellent discriminatory capability across all algorithms. Naive Bayes achieved exceptional performance with an AUC of 0.842 (95% CI: 0.672-1.000), followed closely by Logistic Regression at 0.833 (95% CI: 0.696-0.971) and Random Forest at 0.775 (95% CI: 0.574-0.976). These results indicate that the miRNA biomarker panel is highly predictive of normalization outcomes specifically in female patients. In contrast, male-specific models (*n*=44) showed poor to fair predictive performance. Naive Bayes achieved the highest AUC of 0.617 (95% CI: 0.446-0.787), representing fair discrimination, while both Random Forest (AUC = 0.517, 95% CI: 0.315-0.719) and Logistic Regression (AUC = 0.508, 95% CI: 0.304-0.713) demonstrated poor discriminatory ability with AUCs barely above chance level.

**Figure 3.**
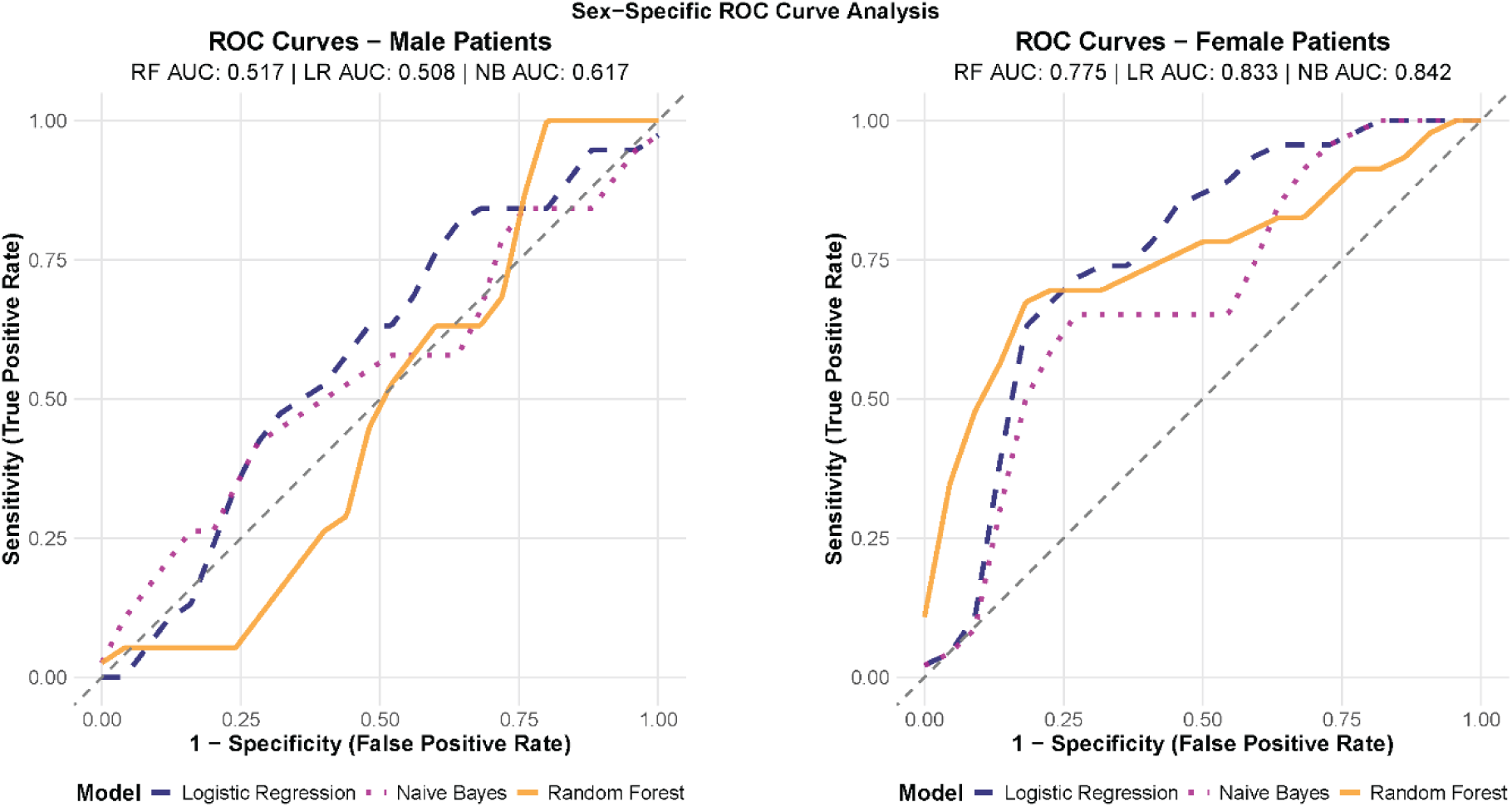
Sex-stratified model performance for normalization prediction. Receiver Operating Characteristic (ROC) curves for Random Forest (RF, yellow), Logistic Regression (LR, dashed-blue), and Naive Bayes (NB, dotted magenta) models stratified by sex using 10-fold cross-validation. Left panel shows male-specific models (n=44); right panel shows female-specific models (n=45). Female-specific models demonstrated superior predictive performance compared to male-specific models. All models used miR-122b-5p, miR-517a-3p, and miR-4683 as predictors. Diagonal dashed line represents random classification (AUC=0.5).

Overall, the 10-fold cross-validation approach provided robust performance estimates with reasonable confidence intervals, particularly for the female-specific models where confidence intervals remained relatively narrow despite smaller sample sizes (n=45). The validation strategy minimized concerns about overfitting while maximizing the use of available data for both training and validation.

#### 3.4.3. Variable Importance Analysis

Variable importance rankings differed markedly across algorithms (Fig. 4A), reflecting complementary predictive mechanisms. Random Forest primarily relied on miR-122b-5p and miR-4683, whereas Logistic Regression emphasized miR-517a-3p. Naive Bayes showed a more balanced contribution from all three biomarkers. These algorithm-dependent importance patterns indicate that the predictive contribution of each miRNA varies according to the modeling framework, reflecting differences in how classifiers exploit linear, nonlinear, and probabilistic structures in the data. This variability underscores the limitations of single-model interpretations and supports the robustness of the consensus based multibiomarker panel identified in this study.

**Figure 4.**
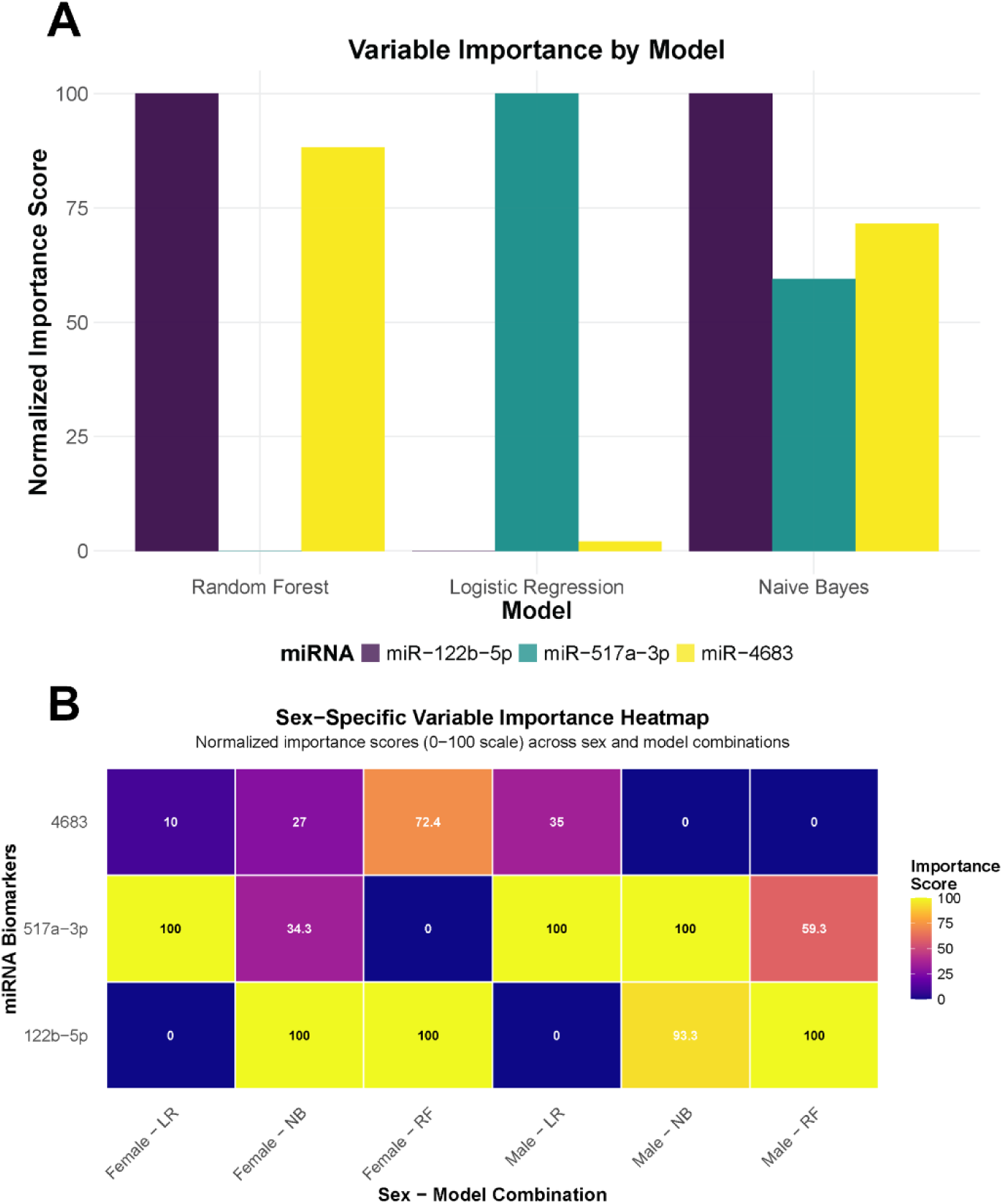
Variable importance of miRNA biomarkers across machine learning global models. **A**: Normalized importance scores (0-100 scale) for miR-122b-5p, miR-517a-3p, and miR-4683 across Random Forest (RF), Logistic Regression (LR), and Naive Bayes (NB) models for the whole AS cohort. **B**: Normalized importance scores (0–100) for miRNA biomarkers across sex-stratified machine learning models. Female-specific models showed superior performance (AUC: 0.775-0.842, 95% CI) with different biomarker ranking patterns compared to male-specific models (AUC: 0.508-0.617, 95% CI). RF = Random Forest, LR = Logistic Regression, NB = Naive Bayes. For brevity, the “hsa-miR” prefix of miRNAs is omitted.

Sex-stratified variable importance analysis revealed differences in the relative contribution of the three miRNA biomarkers between male and female patients (Fig. 4B). In male patients, miR-517a-3p showed the highest and most consistent importance across models, whereas miR-122b-5p and miR-4683 displayed more moderate contributions. In contrast, female patients exhibited a more balanced importance profile, with miR-122b-5p ranking highest, followed by miR-4683 and miR-517a-3p. Consistent with these patterns, statistical significance testing indicated that no miRNA reached significance (p < 0.05) in male patients, while both miR-122b-5p and miR-517a-3p were significantly associated with normalization outcomes in female patients, suggesting more robust biomarker-outcome relationships in this cohort.

### 3.5. Identification and validation of candidate miRNA targets

We focused our analysis on the miRNAs that exhibited the strongest predictive performance during the initial validation phase. Specifically, miR-4683 was excluded from the target analysis, narrowing our scope to those miRNAs that achieved a 100% importance score in at least one sex-specific model (Fig. 4B). This approach allowed us to concentrate on the miRNAs with the greatest statistical robustness and biological significance.

We realized an in-silico analysis of the potential targets for miR-517a-3p and miR-122b. To minimise the number of target genes to analyze, we selected those targets that present the highest score in databases like miRWalk, miRTarBase, and miRDB. We found that miR-517a-3p presented the highest score in the three databases to predict potential binding sites on target drebrin 1 (DBN1) sequence. Drebrin, a developmentally regulated brain protein, has been implicated in critical cellular functions through its interaction with filamentous actin (F-actin), contributing to the modulation of actin filament stability (60). The functional role of drebrin in actin cytoskeletal dynamics has been extensively characterized in neuronal systems (61). However, more recently it has been related with fibrosis-promoting effects in heart and lung (62,63). Therefore, we next assessed the expression levels of DBN1 in our cohort of AS patients and in our experimental mice model of pressure overload subjected to transverse aortic constriction (TAC) and release after 4 weeks TAC (de-TAC) (58).

A linear regression analysis using the expression levels of DBN1 and miR-517a-3p in preoperative LV samples from patients with AS (n=40) revealed an inverse correlation between the two parameters (Fig. 5A). This suggests that, as expected, elevated levels of miR-517a-3p may be silencing the expression of its target gene DBN1 in the myocardium. In addition those patients who did not normalize LV mass 1 year after valve replacement, presented higher levels of preoperative levels of DBN1 compared with patients with good reverse remodelling (Fig. 5B). Moreover, DBN1 LV expression levels were downregulated and upregulated respectively in (male and female) mice under TAC. After 1 week of pressure overload released (de-TAC) the LV levels of DBN1 returned to baseline values (Fig. 5C). Further, we determined DBN1 protein expression levels in the LV of TAC and de-TAC male mice and a clear regulation of the protein levels were observed (Fig. 5D).

**Figure 5.**
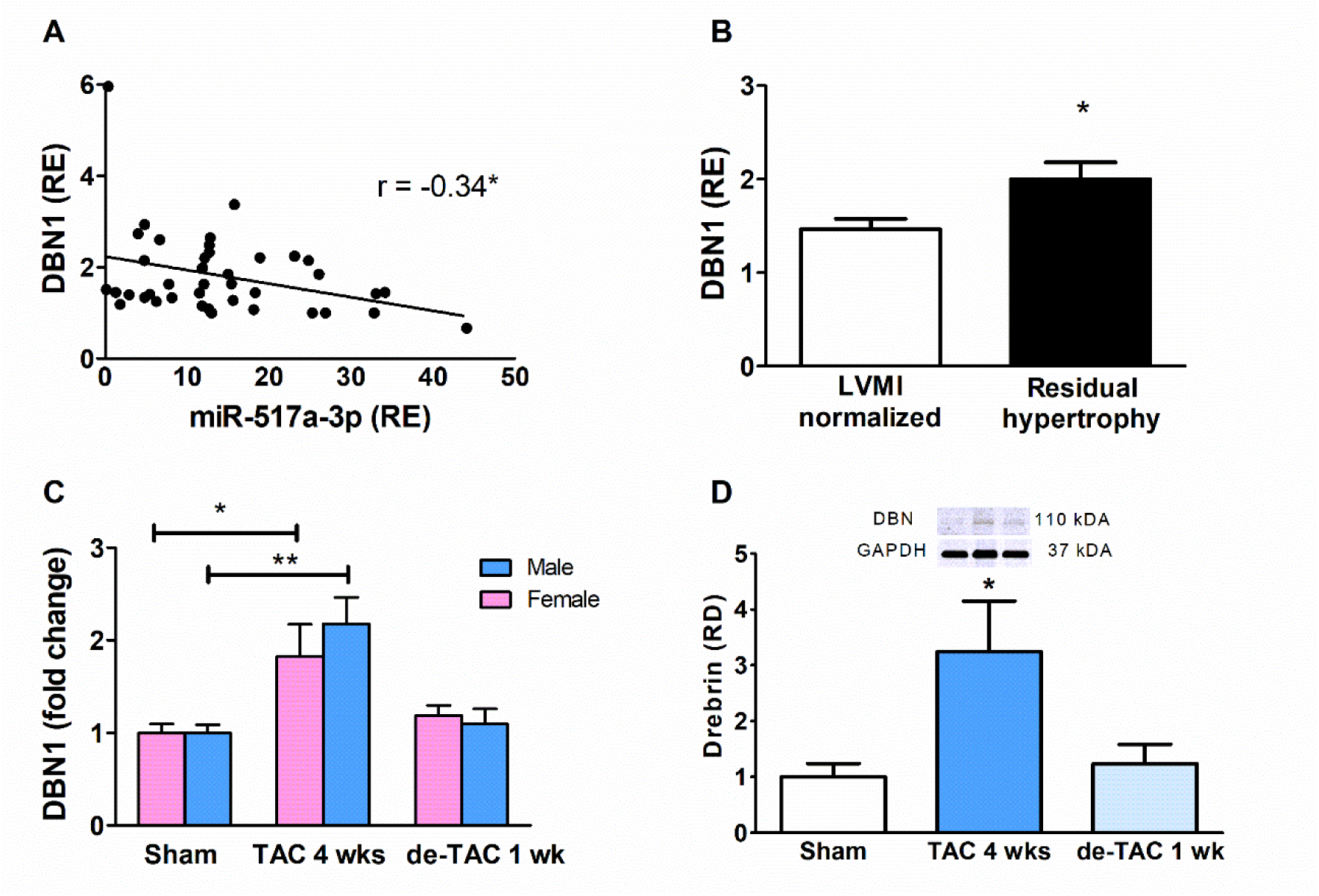
Expression of miR-517a-3p and DBN1 in the LV from AS patients and mice under pressure overload. **A**: Correlation of miR-517a-3p and DBN1 LV relative expression levels (RE vs RNU6b; RE vs 18s) from AS patients (n = 40); r, Pearson correlation coefficient, *p<0.05. **B**: Preoperative levels of DBN1 in LV myocardium in patients who maintained residual hypertrophy left ventricular mass index (LVMI ≥ 51 g/m^2.7^) 1 year after aortic valve replacement and patients who normalized LVMI <51 g/m^2.7^; Student’s t-test, *p<0.05. **C**: Fold change in mRNA levels of DBN1 in the LV from male and female mice subjected to sham surgery, transverse aortic constriction (TAC) and release after 4 weeks TAC (de-TAC); ANOVA followed by the Bonferroni post-hoc test, *p<0.05; **p<0.01. **D**: Representative western blot from male mice (n=4 per group) and average relative density (RD vs GAPDH); ANOVA followed by the Bonferroni post-hoc test, ***p<0.001.

Drebrin is specifically expressed in activated myofibroblasts and enhances actin cytoskeleton assembly, thereby increasing the expression of profibrotic genes such as Acta2 and Col1a1 (23,54). Accordingly, we evaluated the correlation between DBN1 expression and the expression of genes encoding proteins involved in fibrotic processes within the extracellular matrix of the left ventricle in a murine model subjected to pressure overload. Fig. 6 shows a direct correlation between DBN1 and the genes Col1a1 (Fig. 6A), Col1a3 (Fig. 6B), and TGF-β1 (Fig. 6D). Likewise, a direct correlation is also observed between DBN1 and ACTA2 (Fig. 6C), which encodes smooth muscle α-actin (α-SMA), one of the most important markers of myofibroblast activation.

**Figure 6.**
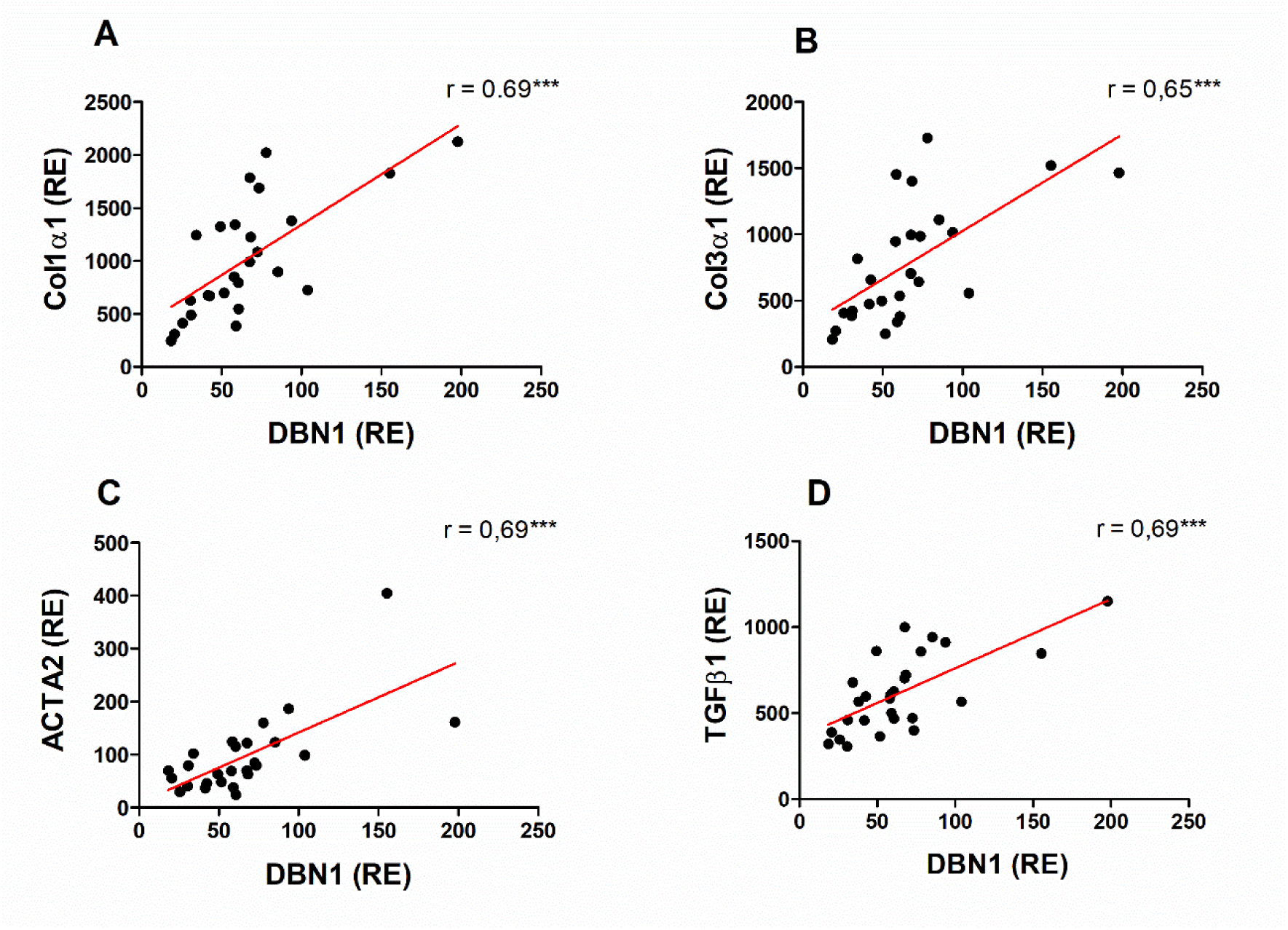
Linear regression and Pearson correlation analysis showing the relationship between the relative expression (RE) of DBN1 and profibrotic genes in the left ventricle of a murine model subjected to pressure overload and relative expression (RE) of A: Col1a1, B: Col1a3, C: ACTA2, and D: TGF-β1. r: Pearson correlation coefficient; **p<0.001.

## 4. DISCUSSION

Aortic valve replacement, performed either surgically or via transcatheter implantation, remains the sole effective intervention for AS and typically initiates reverse cardiac remodelling. However, myocardial recovery is frequently incomplete, and a substantial proportion of patients exhibit persistent left ventricular (LV) hypertrophy after valve replacement, a phenotype associated with impaired long-term outcomes (2,64,65). Moreover, despite extensive investigation, no pharmacological treatment has demonstrated efficacy in preventing or slowing the progression of calcific AS (5). Identifying molecular determinants associated with the reversibility of myocardial remodelling may therefore improve risk stratification and reveal biological pathways potentially amenable to therapeutic modulation. In this context, the integration of transcriptomic profiling with machine learning offers a promising approach for biomarker discovery and for identifying molecular signatures associated with clinically relevant phenotypes (66,67).

Among supervised algorithms, Random Forest is particularly well suited for miRNA profiling due to its ability to handle high-dimensional data, tolerate collinearity, and capture nonlinear relationships (68–71). It also provides variable-importance metrics that facilitate biomarker prioritization. In our analysis, both Mean Decrease Accuracy (MDA) and Mean Decrease Gini (MDG) consistently highlighted miR-4683, miR-122b-5p, and miR-517a-3p as top predictors, and the concordance between these complementary metrics strengthens the robustness of this selection (72,73).

Bayesian Networks offer a complementary framework by modelling probabilistic dependencies and uncovering structural relationships within omics datasets (32,74,75). In settings with many variables and limited sample size, combining structure learning, tabu search, and bootstrap procedures improves network stability and supports the identification of robust Markov Blankets associated with the phenotype of interest (76–78). This strategy has been successfully applied in cardiovascular and neurological disorders to identify relevant biomarkers and integrate complex biological information (79,80). In our study, after 100 bootstrap replications and tabu-based structure learning, the miRNAs most frequently appearing in the Markov Blanket of the node representing left-ventricular mass normalization were miR-122b-5p (39%), miR-4683 (38%), miR-4473 (19%), and miR-517a-3p (17%), supporting the utility of Bayesian Networks for isolating compact sets of variables closely linked to reverse remodelling.

To enhance biomarker robustness, we integrated Random Forest and Bayesian Network outputs through a consensus analysis, combining predictive relevance with probabilistic stability. Hybrid approaches of this type have shown improved biomarker selection in omics studies with limited sample sizes (81,82).The consensus analysis identified miR-4683, miR-122b-5p, and miR-517a-3p as the most robust candidates, given their highest consensus scores (6.6, 6.0, and 3.4, respectively) and strong agreement across methods. In contrast, although miR-4473 appeared relatively frequently in the Bayesian Network, its lower consensus score and limited relevance in Random Forest, particularly in MDA, suggest a weaker association with left-ventricular mass normalization. Overall, the convergence of both analytical strategies increases confidence in the prioritized miRNAs and underscores the value of integrating predictive and probabilistic approaches for transcriptomic analyses in patients with aortic stenosis.

After experimental validation, we assessed, in a cohort of 89 patients, the predictive performance of the three miRNAs for identifying left-ventricular mass normalization one year after valve replacement using three classification algorithms (Random Forest, Naive Bayes, and Logistic Regression). Given the limited sample size, the primary aim was to determine whether the miRNA panel maintained consistent predictive ability across analytical methods. Although Logistic Regression is the most commonly used model for binary clinical outcomes (83), Naive Bayes achieved the highest discriminative performance (AUC = 0.70), in line with previous miRNA-based studies where it has shown robustness under high dimensionality and small sample conditions (84).

One of the most notable findings was the strong sex-specific influence on the predictive performance of the miRNA panel. In women, all three algorithms showed markedly superior discrimination compared with men (AUC in women: RF = 0.78, NB = 0.84, LR = 0.83; versus AUC in men: RF = 0.52, NB = 0.62, LR = 0.51). These results align with existing evidence indicating sex-dependent differences in miRNA regulation and in the molecular mechanisms underlying cardiac remodelling (85–87). Collectively, these data suggest that the molecular determinants of reverse remodelling may differ substantially between women and men, supporting the development of sex-stratified predictive models rather than universal approaches, in line with current precision-medicine strategies (88–90).

The lack of statistical robustness in male patients (with wide 95% confidence intervals for AUC), suggests that the three-biomarker panel may be insufficient for this group, potentially requiring additional biomarkers or alternative molecular targets. Promising candidates such as 4473 or, to a lesser extent– miR-100-5p, which emerged as influential in the exploratory analysis of the sequencing dataset and was consistently supported by RF and BN, warrant further investigation. Other miRNAs, such as miR-342-3p, which were highlighted by RF only, or miR-509-5p by Bayesian networks only, may also play a relevant role. Their limited visibility in the current analysis could be attributed to the small sample size, which may have constrained the detection of subtler associations. A key limitation of this study is the restricted availability of myocardial tissue, which constrained the number of miRNAs that could be experimentally validated. Larger independent cohorts, longitudinal follow-up, and the integration of multi-omics approaches will be required to confirm the clinical utility of these biomarkers and to further elucidate the molecular mechanisms underlying reverse left-ventricular remodelling.

The search for relevant targets focused on those miRNAs that showed the strongest predictive performance during the validation phase (miR-517a-3p and miR-122b-5p). To reduce the number of candidate genes, we selected only those with the highest prediction scores across miRWalk, miRTarBase, and miRDB. miR-517a-3p showed the strongest and most consistent evidence in all three databases, with high-confidence predicted binding sites in the DBN1 sequence.

In contrast, the analysis of miR-122b-5p did not reveal any common high-confidence targets across the three resources.

DBN1 encodes drebrin, an actin-binding protein involved in cytoskeletal organization, cell migration, and actin–microtubule dynamics. Although physiologically expressed mainly in the central nervous system, recent studies show that drebrin is also induced in pathological contexts characterized by intense cytoskeletal remodelling, including tumor progression and fibrosis (60,61,91,92). In hepatic and cardiac fibrosis models, DBN1 upregulation accompanies fibroblast-to-myofibroblast transdifferentiation and promotes F-actin stabilization and collagen production (62,63).

Several findings from our study support the potential relevance of DBN1 to pressure overload-induced myocardial remodelling. First, myocardial DBN1 expression was inversely correlated with miR-517a-3p expression in patients with AS. Second, preoperative DBN1 expression was higher in patients who subsequently failed to normalize LV mass after valve replacement. Third, DBN1 expression was modulated by pressure overload in the murine TAC model and returned towards baseline following removal of the pressure overload. Finally, DBN1 expression correlated positively with several markers associated with fibroblast activation and extracellular matrix remodelling in the murine myocardium. Together, these observations link DBN1 expression to both pressure overload and fibrotic remodelling and support its potential involvement in myocardial adaptation to mechanical stress.

The inverse association between miR-517a-3p and DBN1, together with in silico target prediction, raises the possibility of a miR-517a-3p/DBN1 regulatory axis contributing to myocardial remodelling. Confirmation will require dedicated mechanistic experiments, including modulation of miR-517a-3p expression and direct target-validation assays. Accordingly, the miR-517a-3p/DBN1 relationship should currently be regarded as a biologically plausible regulatory hypothesis emerging from the integration of computational prediction, human myocardial expression data, and experimental pressure-overload models.

Several limitations should be considered when interpreting these findings. First, despite the use of an independent validation cohort, the relatively small sample sizes, particularly after sex stratification, limit the precision of predictive performance estimates. Second, myocardial biopsies provide direct access to disease-relevant tissue but necessarily constrain sample availability and may limit the immediate applicability of the identified miRNAs as clinically accessible biomarkers. Third, the predictive models were based exclusively on molecular variables; future studies should determine whether integration with established clinical and imaging parameters provides incremental predictive value. Finally, although the experimental findings support the biological relevance of DBN1 in pressure overload-induced remodelling, direct regulation by miR-517a-3p and its causal contribution to reverse remodelling remain to be established.

Overall, this study demonstrates the value of integrating myocardial miRNA profiling, complementary machine-learning approaches, and experimental validation to investigate the molecular determinants of reverse cardiac remodelling after aortic valve replacement. Consensus analysis identified a three-miRNA signature associated with subsequent LV mass normalization, with exploratory analyses suggesting potentially greater predictive performance in women. Furthermore, integration of target prediction with human and experimental data identified DBN1 as a biologically plausible candidate linking miR-517a-3p to cytoskeletal and fibrotic remodelling. Although external validation and mechanistic studies are required before clinical translation, these findings provide a framework for identifying molecular signatures associated with myocardial recovery after relief of pressure overload and for exploring regulatory pathways that may ultimately contribute to more individualized management of patients with AS.

## Data Availability

Data will be available within the article or in an appropriate repository at time of publication.

## 6. Acknowledgements

We are grateful to the excellent technical assistance of Nieves García-Iglesias, Beatriz García-Cañón, R.N. and Roberto Moreta, R.N. The authors thank Juan Miguel Cano for his work during the initial phase of the project and for useful exploratory work that informed subsequent analyses.

